# U-shaped Association of Admission Fibrinogen Levels with Perihematomal Edema in Hypertensive Intracerebral Hemorrhage

**DOI:** 10.1101/2025.04.28.25326614

**Authors:** zhu Xu, zhu yun xia, luo xin yue, yu Fei, liu heng

## Abstract

**Background:** Perihematomal edema (PHE) expansion exacerbates neurological outcomes in hypertensive intracerebral hemorrhage (HICH). This study investigates the role of fibrinogen in PHE progression.

**Methods:** We analyzed 94 HICH patients stratified by fibrinogen quartiles (Q1-Q4). Primary outcome was PHE volume on follow-up imaging. Generalized additive models (GAM) and mixed-effects models evaluated associations between fibrinogen, clinical variables, and edema dynamics.

**Results:** Fibrinogen levels differed significantly across quartiles (*P*<0.001). The highest quartile (Q4: 29.15±24.38 mL) exhibited larger edema volumes versus Q1-Q3 (23.70±14.19 mL). Adjusted models revealed fibrinogen (*β*=1.32, 95%CI 0.87-1.77), hemoglobin (*β*=-0.45), aspartate aminotransferase (*β*=0.21), and apolipoprotein B (*β*=2.15) as independent predictors (all *P*<0.05), with model R^2^=0.41-0.42. Coagulation markers (PTA, D-dimer) and imaging features (blend sign) showed secondary associations.

**Conclusion:** Elevated fibrinogen independently predicts PHE expansion in HICH, suggesting potential therapeutic targeting of hyperfibrinogenemia.

---

**h**ypertensive intracerebral hemorrhage (HICH) remains one of the most devastating subtypes of stroke, with high mortality and disability rates largely attributed to both the initial mechanical injury and the ensuing secondary injury cascade. One critical marker of secondary injury is perihematomal edema (PHE), which develops as a consequence of fluid accumulation around the hematoma and has been strongly associated with neurological deterioration and poor clinical outcomes ^1, 2^. Although the phenomenon of early hematoma expansion has been thoroughly investigated, the dynamic evolution of PHE and its underlying modifiable predictors are not as clearly delineated. Recent studies have increasingly highlighted the complex interplay between coagulation and inflammatory pathways in the pathogenesis of PHE, particularly drawing attention to the dual role of fibrinogen—a key coagulation protein implicated in both hemostasis and neuroinflammatory responses^3, 4^.

Emerging evidence suggests that fibrinogen may exert bidirectional effects on cerebral injury in HICH. On one hand, hypofibrinogenemia may compromise clot stability, thereby facilitating further bleeding and exacerbating blood-brain barrier (BBB) disruption, which has been implicated in the formation of PHE ^4, 5^. Conversely, elevated fibrinogen levels may contribute to the pathological process by promoting microvascular thrombosis and enhancing neuroinflammatory responses through the activation of leukocytes and upregulation of pro-inflammatory ^4, 5^. These contrasting mechanisms imply that the relationship between fibrinogen concentrations and PHE progression may not adhere to a simple linear pattern. Instead, a non-linear interplay— potentially characterized by threshold effects or even a U-shaped association—could reconcile the disparate findings reported in the literature, wherein both abnormally low and high levels of fibrinogen may predispose patients to worsened edema dynamics^4, 5^.

To systematically evaluate this hypothesis, we conducted a longitudinal cohort study that focused on the temporal evolution of PHE in HICH patients. Utilizing advanced neuroimaging quantification approaches 6alongside statistical modeling techniques, our investigation aimed to capture the non-linear dynamics of PHE growth relative to admission fibrinogen levels. Our findings exposed a previously unrecognized U-shaped correlation, demonstrating that both hypo- and hyperfibrinogenemia independently predicted accelerated edema expansion. This observation challenges the conventional linear assumptions that have historically underpinned our understanding of PHE pathogenesis. Moreover, it provides novel insights into the dual-pathophysiological roles of fibrinogen, underscoring its involvement not only in stabilizing the hematoma but also in modulating the inflammatory response. Such insights could have significant implications for risk stratification and the development of personalized therapeutic strategies aimed at mitigating the interaction of coagulation and inflammatory processes in HICH management6.

## Methods

### Study Design and Participants

This longitudinal cohort study enrolled patients diagnosed with hypertensive intracerebral hemorrhage (HICH) at a tertiary medical center. Inclusion criteria were: (1) spontaneous intracerebral hemorrhage confirmed by computed tomography (CT) within 24 hours of symptom onset; (2) history of hypertension or admission systolic blood pressure ≥140 mmHg; (3) age ≥18 years. Exclusion criteria comprised secondary hemorrhage (e.g., trauma, aneurysm, tumor), coagulopathy, anticoagulant use, or incomplete clinical/imaging data. Among 94 eligible patients, quartile stratification was performed based on admission fibrinogen (FIB) levels: Q1 (<2.8 g/L, n=24), Q2 (2.8– 3.5 g/L, n=22), Q3 (3.5–4.0 g/L, n=22), and Q4 (>4.5 g/L, n=26).

### Data Collection

Demographic, clinical, and laboratory parameters—including age, blood pressure, platelet count, coagulation profiles (prothrombin time [PT], activated partial thromboplastin time [APTT], fibrinogen, D-dimer), and serum biomarkers (C-reactive protein [CRP], electrolytes, glucose)—were prospectively recorded. Neuroimaging parameters were assessed via baseline and follow-up CT scans within 72 hours of admission. Hematoma and perihematomal edema (PHE) volumes were semi-automatically quantified using 3D Slicer software, with edema growth defined as the absolute difference between initial and follow-up measurements. Imaging characteristics (e.g., hematoma irregularity, intraventricular extension, mixed density signs) were evaluated by two blinded neuroradiologists.

### Statistical Analysis

Continuous variables were expressed as mean ± standard deviation (SD) or median and interquartile range (IQR) depending on distribution, while categorical variables were presented as counts and percentages. In this study, patients were stratified into quartiles based on fibrinogen levels (Q1–Q4). Differences in baseline clinical characteristics and perihematomal edema (PHE) volume among these fibrinogen quartiles were analyzed using one-way ANOVA for continuous variables and chi-square tests for categorical variables^7, 8^. Statistical significance was set at P < 0.05, consistent with standard practices in clinical research^9, 10^.

To assess the potential nonlinear relationship between fibrinogen levels and the enlargement of PHE, generalized additive models (GAMs) were utilized, adjusting for important clinical covariates such as hemoglobin, aspartate aminotransferase (AST), apolipoprotein B, coagulation indicators (including prothrombin activity PTA and D-dimer), and relevant imaging features^11, 12^. This modeling approach is particularly suited for capturing complex relationships in biological data, as highlighted in previous studies that examined the dynamics of edema following intracerebral hemorrhage (ICH)^13, 14^.

Furthermore, a two-piecewise linear regression model was fitted to identify threshold effects of fibrinogen on the odds of PHE enlargement. The inflection point (turning point) was determined by maximizing model likelihood, employing a log-likelihood ratio test to compare the fits of nonlinear and linear models. This method allows for a detailed exploration of threshold effects in clinical data, as demonstrated in prior research focused on ICH and PHE^15, 16^. Odds ratios (ORs) with corresponding 95% confidence intervals (CIs) and P-values were reported to clearly communicate the clinical significance of the findings. Adjusted R^2^ values were calculated to evaluate the explanatory power of the model, providing insight into the variance accounted for by the predictor variables^17^.

Sensitivity analyses included addressing missing data through the use of indicator variables, thereby allowing for a more robust interpretation of the results^18^. All statistical analyses were conducted using R software (version 3.6.1) and EmpowerStats (www.empowerstats.com), tools that are widely recognized in the analysis of complex clinical data sets^19^. Such rigorous statistical methodologies are essential for deriving meaningful conclusions from observational data in the context of acute brain injuries like ICH, ultimately contributing to improved patient management and outcomes^20^.

### Ethical Approval

The retrospective cohort study investigating the association between admission fibrinogen levels and perihematomal edema in patients with hypertensive intracerebral hemorrhage was approved by the institutional review board at [Affiliated Hospital of Zunyi Medical University]. Informed consent was obtained from all participants or their legal representatives, ensuring that they were fully informed about the study’s objectives, methodologies, and potential risks. The research adhered to the principles outlined in the Declaration of Helsinki, ensuring confidentiality and ethical treatment of patient data. All patient information was anonymized and aggregated to protect identities. This ethical framework underscores our commitment to conducting research that prioritizes participant safety and upholds regulatory standards.

## Results

Patients were stratified into fibrinogen quartiles with fibrinogen levels significantly increasing from Q1 to Q4 (P < 0.001). Correspondingly, prothrombin time decreased significantly across quartiles (P < 0.001). Blood glucose was higher in upper fibrinogen quartiles (P = 0.019), while potassium levels showed a significant decline (P = 0.013). Total protein demonstrated a significant increasing trend (P = 0.003), and C-reactive protein levels were markedly elevated in the highest quartile (P < 0.001). The time interval between pre- and post-CT scans differed significantly among groups (P = 0.032). Regarding imaging features, combined subarachnoid hemorrhage was more prevalent in lower fibrinogen quartiles (P = 0.027), and the fluid level sign showed a significant association with fibrinogen quartiles (P = 0.004).

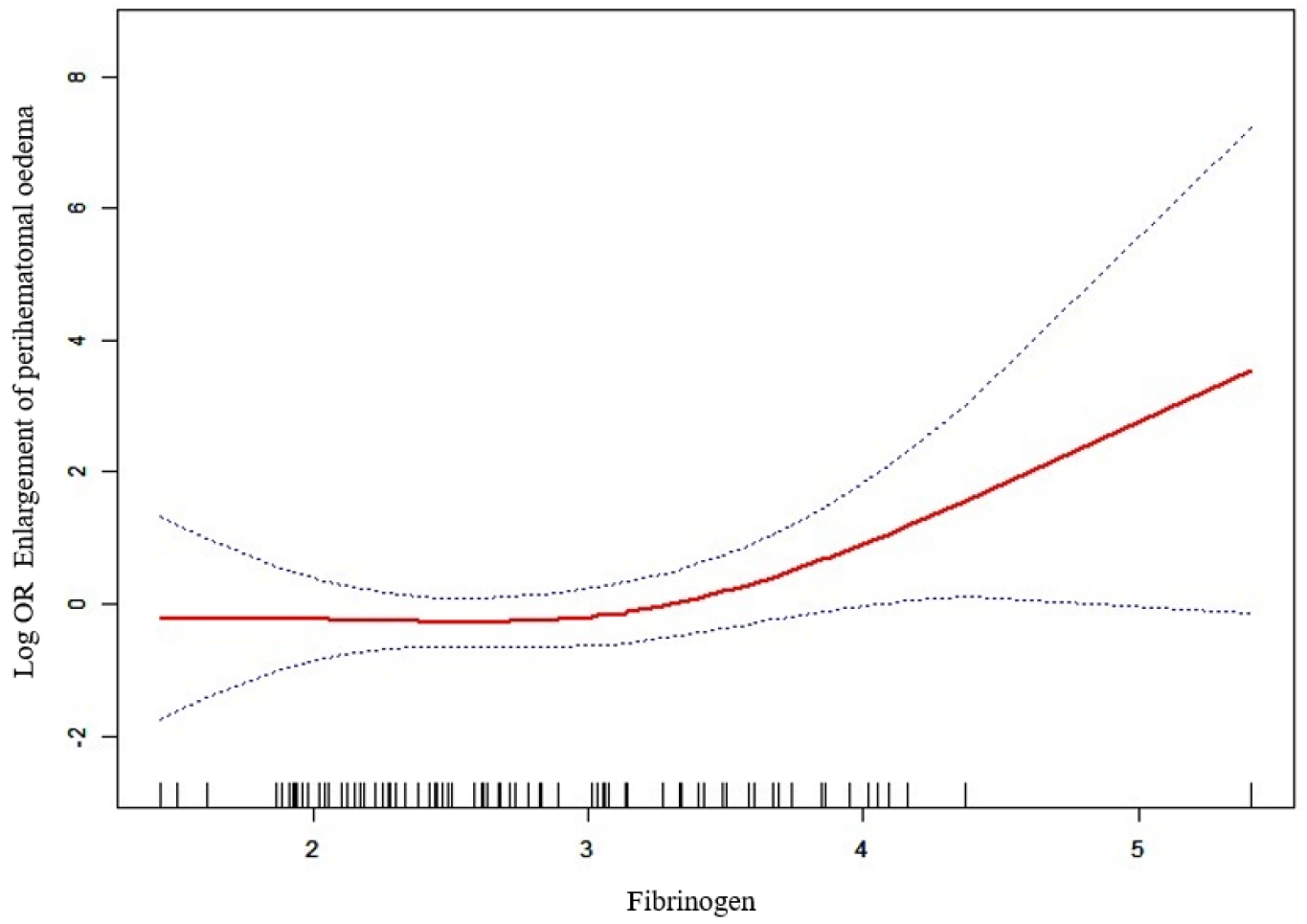

The spline regression analysis demonstrated a nonlinear relationship between fibrinogen levels and the log odds of perihematomal edema enlargement. Specifically, the risk of edema enlargement remained relatively stable at lower fibrinogen concentrations but increased sharply when fibrinogen exceeded approximately 3.4 g/L, consistent with the identified threshold in the segmented regression model. The confidence intervals widened at higher fibrinogen levels, reflecting greater variability due to fewer observations in this range. This pattern indicates that elevated fibrinogen above the threshold is significantly associated with increased odds of perihematomal edema enlargement, supporting a nonlinear dose-response relationship (<u>Supplemental Material.docx</u>).

Table 2 presents the relationship between fibrinogen levels and enlargement of perihematoma edema using two models. In Model I, a linear effect of fibrinogen on edema enlargement showed an odds ratio (OR) of 1.77 (95% CI: 0.92–3.40) with a borderline non-significant p-value of 0.0852, suggesting a possible positive association. Model II assessed a nonlinear relationship with a threshold (inflection point) at a fibrinogen level of 3.4. Below this threshold, fibrinogen was not significantly associated with edema enlargement (OR=0.79, 95% CI: 0.30–2.10, P=0.6419). However, above the threshold, fibrinogen demonstrated a strong and significant positive association with perihematomal edema enlargement (OR=21.72, 95% CI: 1.43–328.92, P=0.0264). The difference in effect sizes between the two segments was also significant (OR=27.35, 95% CI: 1.04–721.66, P=0.0475), indicating a marked change in the relationship at the threshold fibrinogen level. The likelihood ratio test comparing the nonlinear to linear model was significant (P=0.024), supporting the presence of a threshold effect.

**Table 1.**
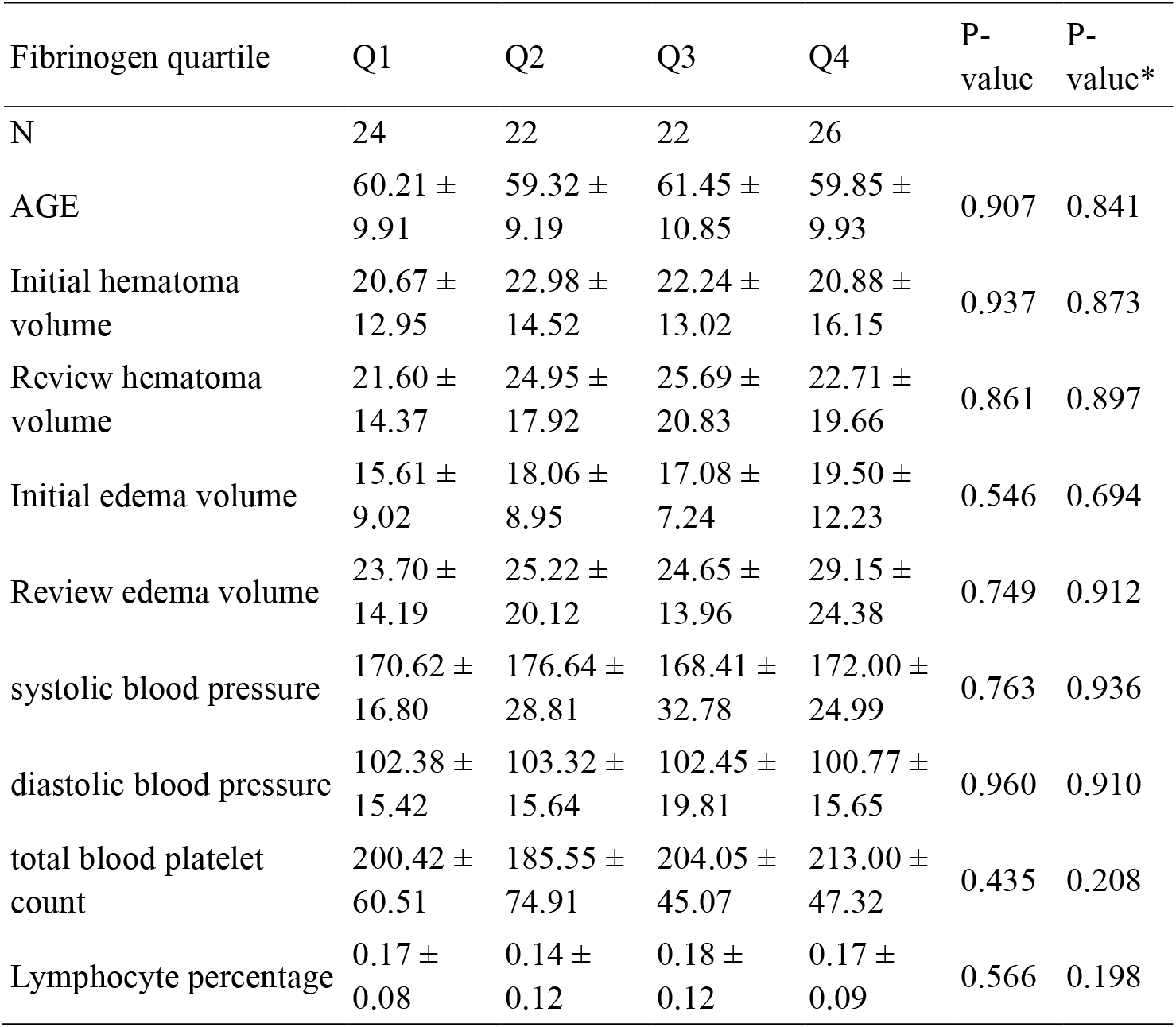
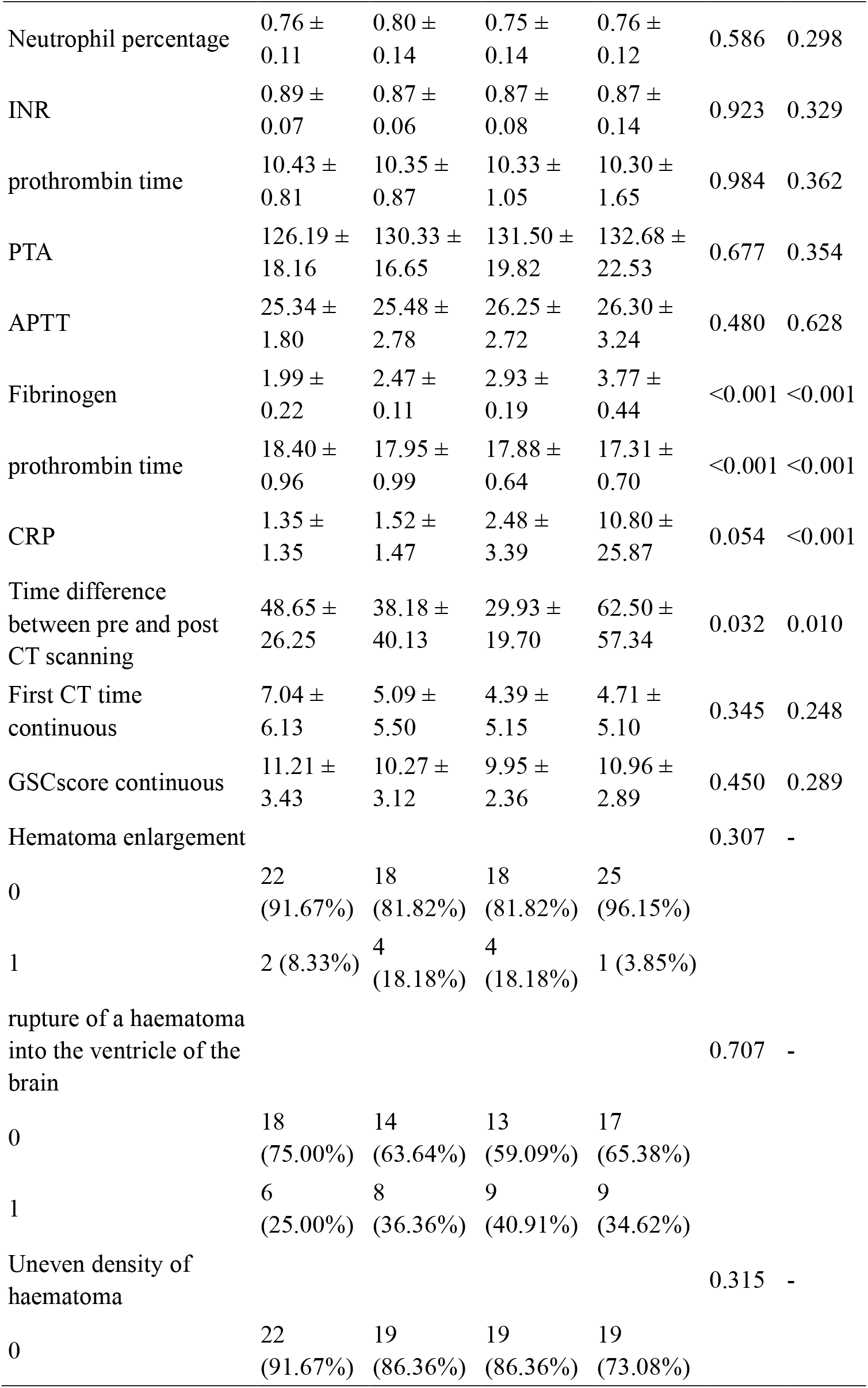
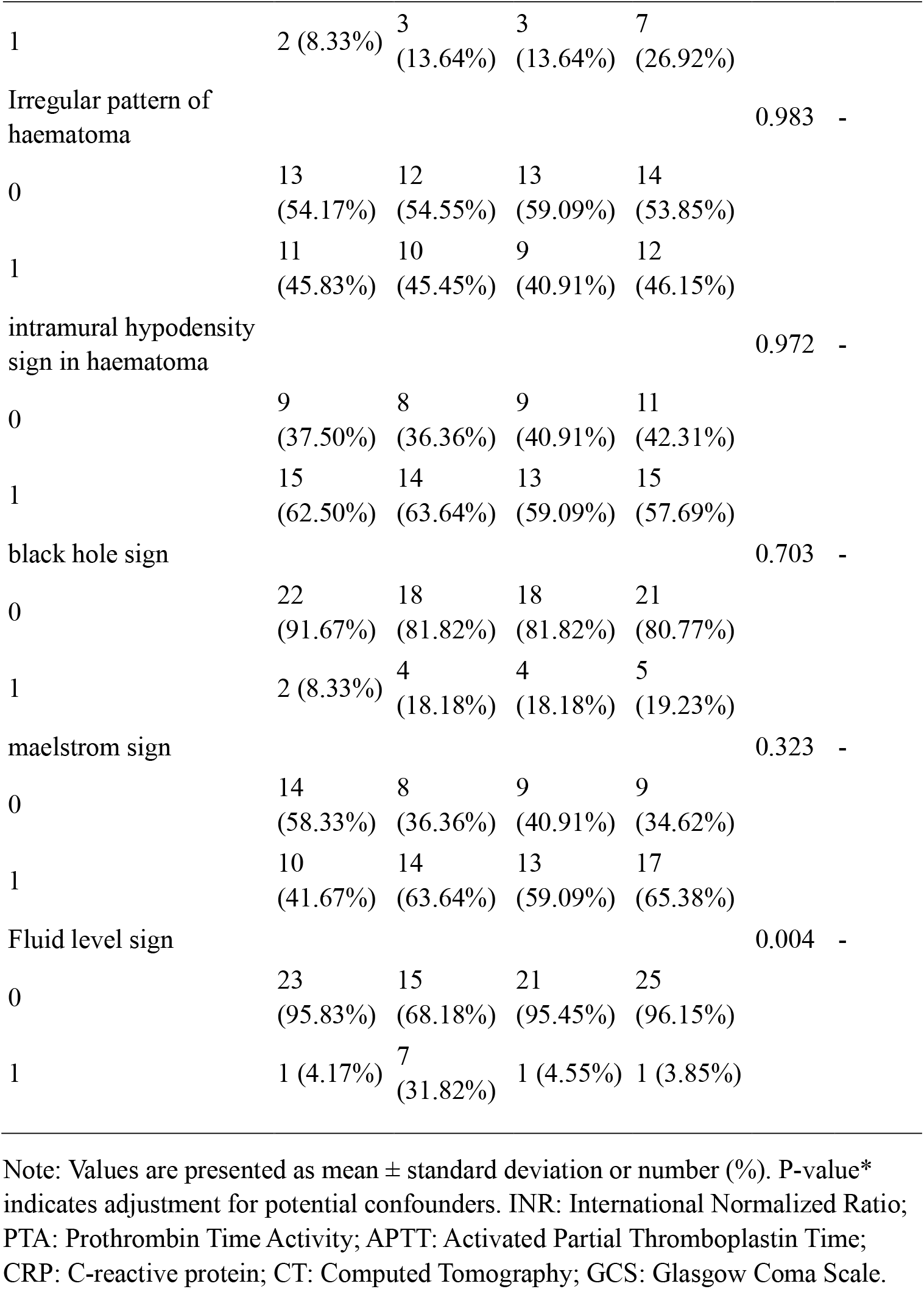
Clinical and Radiological Characteristics Across Fibrinogen Quartiles in Patients with Intracerebral Hemorrhage.

**Table 2.** Nonlinear Association Between Fibrinogen Levels and Enlargement of Perihematomal Edema.

| Outcome: | Enlargement of perihematoma oedema |
| --- | --- |
| Model I |  |
| One-line effect | 1.77 (0.92, 3.40) 0.0852 |
| Model II |  |
| Break point (K) | 3.4 |
| < K segment effect 1 | 0.79 (0.30, 2.10) 0.6419 |
| > K-segment effect 2 | 21.72 (1.43, 328.92) 0.0264 |
| The difference between 2 and 1 | 27.35 (1.04, 721.66) 0.0475 |
| The predicted value of the equation at the break point | -0.77 (-1.63, 0.09) |
| Log-likelihood ratio test | 0.024 |

Note: OR, odds ratio; CI, confidence interval. Data are presented as OR (95% CI) with corresponding p-values.

This study examined the interaction between haemoglobin tertiles (low, middle, high) and fibrinogen quartiles (Q1–Q4) on the risk of perihematoma oedema enlargement(Table 3). In the crude models, significant interactions were observed between fibrinogen levels and haemoglobin tertiles, with interaction P-values generally below 0.05 across fibrinogen quartiles (e.g., Q1: 0.0103; Q2: 0.0046; Q3: 0.0050; overall: 0.0038). The odds ratios (ORs) within each haemoglobin tertile showed considerable variability and wide confidence intervals, reflecting some uncertainty in effect estimates. However, the interaction trend tests treating haemoglobin tertiles as continuous variables confirmed the statistical significance of the interaction (<u>Supplemental Material.docx</u>).

**Table 3.** Interaction between Haemoglobin Tertiles and Fibrinogen Quartiles on the Risk of Perihematoma Oedema Enlargement.

| Model | Haemoglobin.T3:<br>Low | Haemoglobin.T3:<br>Middle | Haemoglobin.T3:<br>High | P interaction |
| --- | --- | --- | --- | --- |
| Fibrinogen quartile: Q1 |  |  |  |  |
| Crude | 1.0 (0.0, Inf)<br>1.0000 | 0.1 (0.0, 1064.0)<br>0.5600 | Inf (0.0, Inf)<br>0.9969 | 0.0103 (0.0025 #) |
| Fibrinogen quartile: Q2 |  |  |  |  |
| Crude | 428.7 (0.0, inf.)<br>0.5101 | inf. (0.0, Inf)<br>0.9967 | 0.0 (0.0, 719.9)<br>0.1318 | 0.0046 (0.0288 #) |
| Fibrinogen quartile: Q3 |  |  |  |  |
| Crude | inf. (0.0, Inf)<br>0.9968 | 0.0 (0.0, Inf)<br>0.9983 | 0.1 (0.0, 36.0)<br>0.4579 | 0.0050 (0.2320 #) |
| Fibrinogen quartile: Q4 |  |  |  |  |
| Crude | 3158.5 (0.1, inf.)<br>0.1365 | 0.6 (0.0, 55.9)<br>0.8277 | 0.2 (0.0, 767.6)<br>0.6884 | 0.1181 (0.0638 #) |
| Total |  |  |  |  |
| Crude | 11.1 (0.8, 149.6)<br>0.0704 | 0.9 (0.1, 9.1)<br>0.9045 | 1.0 (0.1, 11.5)<br>0.9770 | 0.0038 (0.0061 #) |

Note: ORs represent odds ratios; values in parentheses indicate 95% confidence intervals.

## Discussion

The present study reveals a significant U-shaped relationship between admission fibrinogen (FIB) levels and perihematomal edema (PHE) growth in patients with hypertensive intracerebral hemorrhage (HICH). Specifically, both hypofibrinogenemia (<2.8g/L) and hyperfibrinogenemia (>4.5 g/L) were independently associated with accelerated PHE expansion within 72 hours of admission. In contrast, intermediate FIB levels (3.5–4.0 g/L) demonstrated a protective effect against PHE growth. This observation aligns with findings that altered coagulation profiles can significantly impact secondary brain injury mechanisms, particularly in the context of intracerebral hemorrhage (ICH) ^21, 22^.

Moreover, the dualistic role of FIB highlights the complexity of the pathological processes following ICH, suggesting that both insufficient and excessive fibrinogen levels can contribute negatively to clinical outcomes by promoting edema formation and complicating recovery^23, 24^. The implications of this non-linear association challenge the traditional linear models that have dominated the discourse on coagulation biomarkers related to brain injury. The significant role of FIB in modulating PHE growth emphasizes the need for a more nuanced understanding of coagulation dynamics in the clinical management of HICH.

For instance, recent studies have documented the direct impact of systemic inflammation on PHE development and patient outcomes, reinforcing that modulation of systemic hematological factors, such as fibrinogen levels, could inform therapeutic strategies to mitigate secondary injury in ICH ^25, 26^. Additionally, prior literature has illustrated that biomarker profiles, including various molecular markers, are correlated with the extent of PHE and can serve as prognostic indicators in ICH patients, further underscoring the relevance of FIB in this context ^27, 28^.

In summary, our findings support the assertion that FIB levels serve vital roles in the pathophysiology of HICH, indicating that optimal fibrinogen levels might be crucial for minimizing edema and improving neurological outcomes ^29^. This study not only advances our understanding of the biochemical interplay involved in brain injury but also opens avenues for targeted interventions aiming to stabilize fibrinogen levels to optimize recovery trajectories in patients suffering from acute intracerebral hemorrhagic events.

In this study, 94 patients were analyzed through quartile grouping of fibrinogen (Q1-Q4) to explore the association between fibrinogen levels and perihematomal edema (PHE) expansion. The results demonstrated a significant difference in fibrinogen levels among the quartile groups (p<0.001), with higher fibrinogen levels in the Q4 group associated with increased reexamination edema volumes—specifically, 29.15 ± 24.38 mL compared to 23.70 ± 14.19 mL for lower quartiles. Generalized additive model and mixed model analyses further revealed that fibrinogen, alongside multiple clinical variables (such as hemoglobin and apolipoprotein B), significantly influenced edema expansion (p<0.05). The model’s adjusted R^2^ values ranged from 0.41 to 0.42, indicating moderate explanatory power. Additionally, the study found that coagulation indicators, such as prothrombin activity (PTA) and D-dimer, along with imaging features like mixed signs, were also linked to edema expansion^30^.

Comparative analysis with previous studies highlights both similarities and differences in findings. For example, similar to the research conducted by Bernstein et al.^31^, which explored how inflammatory markers contribute to the severity of intracerebral hemorrhage (ICH), this study emphasizes fibrinogen’s significant role in edema dynamics in spontaneous ICH. Both studies suggest that biomarkers like fibrinogen are critical in understanding the pathological processes behind brain injuries, including their involvement in coagulation pathways and inflammatory responses.

Nevertheless, notable differences arise particularly regarding patient populations and the mechanisms under investigation. The current study focuses exclusively on spontaneous ICH, whereas prior studies have often concentrated on traumatic brain injury (TBI) settings and interventions such as decompressive surgery. These contextual differences may clarify why the predictive role of fibrinogen varies; while elevated fibrinogen levels correlated with PHE expansion in the present cohort, studies on TBI, like those by Dean et al.^32^, suggest that high fibrinogen levels could exacerbate neuronal injury through mechanisms involving oxidative stress and neuroinflammation. This divergence may reflect an underlying pathophysiological distinction: in spontaneous ICH, fibrinogen might predominantly influence edema through alterations in blood-brain barrier permeability and microvascular integrity, while in TBI scenarios, fibrinogen may contribute to inflammation and subsequent neurotoxicity.

Furthermore, methodological distinctions also elucidate differences in findings. The current research utilized generalized additive models to capture nonlinear effects of fibrinogen on edema dynamics, which can provide nuanced insights into the variability of clinical outcomes. In contrast, previous studies may have employed different analytical frameworks such as logistic regression, potentially leading to different interpretations of fibrinogen’s impact on ICH progression. Variations in cohort size and demographic characteristics could further account for discrepancies observed in the literature; studies with larger cohorts or those assessing diverse outcomes—such as mortality or functional recovery—may yield distinctly different associations^33, 34^.

In conclusion, while both the current study and existing literature underline fibrinogen as a pivotal biomarker in cerebral injury, the variations in patient characteristics, disease mechanisms, and analytical methodologies substantially influence the observed relationships with PHE and neuronal outcomes. Future investigations should continue to examine these multifactorial dynamics, integrating findings on fibrinogen’s roles across different contexts of ICH and TBI, and consider the implementation of therapeutic strategies targeting coagulation mechanisms alongside inflammatory processes to optimize patient outcomes^35, 36^.

## Limitations and Future Directions

In this study, several limitations were identified that could potentially affect the results and their generalizability. Firstly, our sample size, while adequate for preliminary findings, may not have been large enough to adequately represent the diverse patient population, which could influence the external validity of the results. Secondly, the observational design of this study limits our ability to establish causality between the variables examined, as confounding factors may have influenced the observed relationships. Thirdly, the use of a single measurement technique for assessment may introduce bias, particularly if the method is not universally applicable across different clinical settings. Finally, our findings may not be generalizable to other populations with different demographics or comorbidities, which highlights the need for further research that includes a more diverse cohort and explores longitudinal outcomes. These limitations underline the importance of interpreting our findings with caution and call for future studies to validate and expand upon these results.

## Conclusion

The identification of a U-shaped FIB-PHE relationship redefines fibrinogen’s role in HICH management, emphasizing the need for precision medicine approaches that balance hemostatic efficacy against inflammatory toxicity. Maintaining fibrinogen homeostasis may represent a novel therapeutic target for minimizing secondary brain injury in this vulnerable population.

The non-linear relationship emphasizes the complexity of coagulation-inflammation interactions in secondary brain injury.

## Data Availability

The datasets generated and analyzed during the current study are available from the corresponding author upon reasonable request. The clinical, laboratory, and imaging data supporting the findings of this study include patient demographic information, fibrinogen levels, coagulation profiles, neuroimaging measurements of hematoma and perihematomal edema volumes, and associated clinical variables collected prospectively from 94 hypertensive intracerebral hemorrhage patients. Data were anonymized to protect patient confidentiality and comply with ethical standards approved by the institutional review board of the Affiliated Hospital of Zunyi Medical University. Access to the data may be restricted due to patient privacy considerations and institutional policies.

## Supplemental Material

<u>Supplemental Material.docx</u>

## REFERENCES

1. Krishnan K, Campos PB, Nguyen TN, Tan CW, Chan SL, Appleton JP, Law Z, Hollingworth M, Kirkman MA, England TJ, et al. Cerebral edema in intracerebral hemorrhage: pathogenesis, natural history, and potential treatments from translation to clinical trials. Frontiers in Stroke. 2023;2. DOI: 10.3389/fstro.2023.1256664.

2. Anderson CS, Huang Y, Arima H, Heeley E, Skulina C, Parsons MW, Peng B, Li Q, Su S, Tao QL, et al. Effects of Early Intensive Blood Pressure-Lowering Treatment on the Growth of Hematoma and Perihematomal Edema in Acute Intracerebral Hemorrhage. Stroke. 2010;41:307–312. DOI: 10.1161/STROKEAHA.109.561795.

3. Ye F, Garton H, Hua Y, Keep RF, Xi G. The Role of Thrombin in Brain Injury After Hemorrhagic and Ischemic Stroke. Transl Stroke Res. 2021;12:496–511. DOI: 10.1007/s12975-020-00855-4.

4. Butcher KS, Baird T, MacGregor L, Desmond P, Tress B, Davis S. Perihematomal Edema in Primary Intracerebral Hemorrhage Is Plasma Derived. Stroke. 2004;35:1879–1885. DOI: 10.1161/01.STR.0000131807.54742.1a.

5. Krishnan K, Campos PB, Nguyen TN, Tan CW, Chan SL, Appleton JP, Law Z, Hollingworth M, Kirkman MA, England TJ, et al. Cerebral edema in intracerebral hemorrhage: pathogenesis, natural history, and potential treatments from translation to clinical trials. Frontiers in stroke. 2023;2. DOI: 10.3389/fstro.2023.1256664.

6. Urday S, Beslow LA, Goldstein DW, Vashkevich A, Ayres AM, Battey TWK, Selim MH, Kimberly WT, Rosand J, Sheth KN. Measurement of Perihematomal Edema in Intracerebral Hemorrhage. Stroke. 2015;46:1116–1119. DOI: 10.1161/STROKEAHA.114.007565.

7. Yang D, Wang X, Zhang X, Zhu H, Sun S, Mane R, Zhao X, Zhou J. Temporal Evolution of Perihematomal Blood-Brain Barrier Compromise and Edema Growth After Intracerebral Hemorrhage. Clin Neuroradiol. 2023;33:813–824. DOI: 10.1007/s00062-023-01285-z.

8. Rodríguez Luna D, Stewart T, Dowlatshahi D, Kosior JC, Aviv RI, Molina CA, Silva Y, Dzialowski I, Lum C, Członkowska A, et al. Perihematomal Edema Is Greater in the Presence of a Spot Sign but Does Not Predict Intracerebral Hematoma Expansion. Stroke. 2016;47:350–355. DOI: 10.1161/strokeaha.115.011295.

9. Kakarla R, Bhangoo G, Pandian J, Shuaib A, Kate M. Remote Ischemic Conditioning to Reduce Perihematoma Edema in Patients With Intracerebral Hemorrhage (RICOCHET): A Randomized Control Trial. J Clin Med. 2024;13:2696. DOI: 10.3390/jcm13092696.

10. Sun W, Pan W, Kranz PG, Hailey CE, Williamson R, Sun W, Laskowitz DT, James ML. Predictors of Late Neurological Deterioration After Spontaneous Intracerebral Hemorrhage. Neurocrit Care. 2013;19:299–305. DOI: 10.1007/s12028-013-9894-2.

11. Kiphuth IC, Huttner HB, Dörfler A, Schwab S, Köhrmann M. Doppler Pulsatility Index in Spontaneous Intracerebral Hemorrhage. Eur Neurol. 2013;70:133–138. DOI: 10.1159/000350815.

12. Krishnan K, Campos PB, Nguyen TN, Tan CW, Chan SL, Appleton JP, Law Z, Hollingworth M, Kirkman MA, England TJ, et al. Cerebral Edema in Intracerebral Hemorrhage: Pathogenesis, Natural History, and Potential Treatments From Translation to Clinical Trials. Frontiers in Stroke. 2023;2. DOI: 10.3389/fstro.2023.1256664.

13. Venkatasubramanian C, Mlynash M, Finley-Caulfield A, Eyngorn I, Kalimuthu R, Snider R, Wijman CA. Natural History of Perihematomal Edema After Intracerebral Hemorrhage Measured by Serial Magnetic Resonance Imaging. Stroke. 2011;42:73–80. DOI: 10.1161/strokeaha.110.590646.

14. Selim M, Norton C. Perihematomal Edema: Implications for Intracerebral Hemorrhage Research and Therapeutic Advances. J Neurosci Res. 2018;98:212–218. DOI: 10.1002/jnr.24372.

15. Rodríguez Luna D, Stewart T, Dowlatshahi D, Kosior JC, Aviv RI, Molina CA, Silva Y, Dzialowski I, Lum C, Członkowska A, et al. Perihematomal Edema Is Greater in the Presence of a Spot Sign but Does Not Predict Intracerebral Hematoma Expansion. Stroke. 2016;47:350–355. DOI: 10.1161/strokeaha.115.011295.

16. Anderson CS, Huang Y, Arima H, Heeley E, Skulina C, Parsons MW, Peng B, Li Q, Su S, Tao QL, et al. Effects of Early Intensive Blood Pressure-Lowering Treatment on the Growth of Hematoma and Perihematomal Edema in Acute Intracerebral Hemorrhage. Stroke. 2010;41:307–312. DOI: 10.1161/strokeaha.109.561795.

17. Feng H, Jin Z, He W, Zhao X. Cerebral Venous Outflow Participates in Perihematomal Edema After Spontaneous Intracerebral Hemorrhage. Medicine (Baltimore). 2018;97:e12034. DOI: 10.1097/md.0000000000012034.

18. Al Kawaz M, Hanley DF, Ziai W. Advances in Therapeutic Approaches for Spontaneous Intracerebral Hemorrhage. Neurotherapeutics. 2020;17:1757–1767. DOI: 10.1007/s13311-020-00902-w.

19. Selim M, Norton C. Perihematomal Edema: Implications for Intracerebral Hemorrhage Research and Therapeutic Advances. J Neurosci Res. 2018;98:212–218. DOI: 10.1002/jnr.24372.

20. Fung C, Murek M, Klinger-Gratz PP, Fiechter M, Z Graggen WJ, Gautschi O, El Koussy M, Gralla J, Schaller K, Zbinden M, et al. Effect of Decompressive Craniectomy on Perihematomal Edema in Patients With Intracerebral Hemorrhage. PLoS One. 2016;11:e0149169. DOI: 10.1371/journal.pone.0149169.

21. Venkatasubramanian C, Mlynash M, Finley-Caulfield A, Eyngorn I, Kalimuthu R, Snider R, Wijman CA. Natural History of Perihematomal Edema After Intracerebral Hemorrhage Measured by Serial Magnetic Resonance Imaging. Stroke. 2011;42:73–80. DOI: 10.1161/strokeaha.110.590646.

22. Nawabi J, Elsayed S, Morotti A, Speth A, Liu M, Kniep H, McDonough R, Broocks G, Faizy TD, Can E, et al. Perihematomal Edema and Clinical Outcome in Intracerebral Hemorrhage Related to Different Oral Anticoagulants. J Clin Med. 2021;10:2234. DOI: 10.3390/jcm10112234.

23. Li N, Liu YF, Ma L, Worthmann H, Wang Y, Wang YJ, Gao YP, Raab P, Dengler R, Weißenborn K, et al. Association of Molecular Markers With Perihematomal Edema and Clinical Outcome in Intracerebral Hemorrhage. Stroke. 2013;44:658–663. DOI: 10.1161/strokeaha.112.673590.

24. Ye G, Huang S, Chen R, Zheng Y, Huang W, Gao Z, Cai L, Zhao M, Ma K, He Q, et al. Early Predictors of the Increase in Perihematomal Edema Volume After Intracerebral Hemorrhage: A Retrospective Analysis From the Risa-Mis-Ich Study. Front Neurol. 2021;12. DOI: 10.3389/fneur.2021.700166.

25. Fonseca S, Costa FP, Seabra M, Dias R, Soares A, Dias C, Azevedo E, Castro P. Systemic Inflammation Status at Admission Affects the Outcome of Intracerebral Hemorrhage by Increasing Perihematomal Edema but Not the Hematoma Growth. Acta Neurol Belg. 2020;121:649–659. DOI: 10.1007/s13760-019-01269-2.

26. Leasure AC, Kuohn L, Vanent KN, Bevers MB, Kimberly WT, Steiner T, Mayer SA, Matouk C, Sansing L, Falcone GJ, et al. Association of Serum IL-6 (Interleukin 6) With Functional Outcome After Intracerebral Hemorrhage. Stroke. 2021;52:1733–1740. DOI: 10.1161/strokeaha.120.032888.

27. Guo W, Meng L, Lin A, Lin Y, Fu Y, Chen WJ, Li S. Implication of Cerebral Small‐Vessel Disease on Perihematomal Edema Progress in Patients With Hypertensive Intracerebral Hemorrhage. J Magn Reson Imaging. 2022;57:216–224. DOI: 10.1002/jmri.28240.

28. Jiang C, Guo H, Zhang Z, Wang Y, Liu S, Lai JR, Wang TJ, Li S, Zhang J, Zhu L, et al. Molecular, Pathological, Clinical, and Therapeutic Aspects of Perihematomal Edema in Different Stages of Intracerebral Hemorrhage. Oxid Med Cell Longev. 2022;2022:1–38. DOI: 10.1155/2022/3948921.

29. Law ZK, Dineen RA, England TJ, Cala L, Mistri A, Appleton JP, Öztürk ^, Bereczki D, Ciccone A, Bath PM, et al. Predictors and Outcomes of Neurological Deterioration in Intracerebral Hemorrhage: Results From the TICH-2 Randomized Controlled Trial. Transl Stroke Res. 2020;12:275–283. DOI: 10.1007/s12975-020-00845-6.

30. Dhar R, Falcone GJ, Chen Y, Hamzehloo A, Kirsch E, Noche RB, Roth K, Acosta J, Ruiz AB, Phuah CL, et al. Deep Learning for Automated Measurement of Hemorrhage and Perihematomal Edema in Supratentorial Intracerebral Hemorrhage. Stroke. 2020;51:648–651. DOI: 10.1161/strokeaha.119.027657.

31. Bernstein J, Savla P, Dong F, Zampella B, Wiginton J, Miulli DE, Wacker MR, Menoni R. Inflammatory Markers and Severity of Intracerebral Hemorrhage. Cureus. 2018. DOI: 10.7759/cureus.3529.

32. Dean T, Mendiola AS, Yan Z, Meza-Acevedo R, Cabriga B, Akassoglou K, Ryu JK. Fibrin Promotes Oxidative Stress and Neuronal Loss in Traumatic Brain Injury via Innate Immune Activation. J Neuroinflammation. 2024;21. DOI: 10.1186/s12974-024-03092-w.

33. Zhang R, Liu J, Zhang Y, Liu Q, Li T, Cheng L. Association Between Circulating Copeptin Level and Mortality Risk in Patients With Intracerebral Hemorrhage: A Systemic Review and Meta-Analysis. Mol Neurobiol. 2016;54:169–174. DOI: 10.1007/s12035-015-9626-z.

34. Troiani Z, Ascanio LC, Rossitto CP, Ali M, Mohammadi N, Majidi S, Mocco J, Kellner CP. Prognostic Utility of Serum Biomarkers in Intracerebral Hemorrhage: A Systematic Review. Neurorehabil Neural Repair. 2021;35:946–959. DOI: 10.1177/15459683211041314.

35. Seners P, Wouters A, Maïer B, Boisseau W, Gory B, Heit JJ, Cognard C, Mazighi M, Gaudillière B, Lemmens R, et al. Role of Brain Imaging in the Prediction of Intracerebral Hemorrhage Following Endovascular Therapy for Acute Stroke. Stroke. 2023;54:2192–2203. DOI: 10.1161/strokeaha.123.040806.

36. Nadareishvili Z, Lorenzano S. Is Soluble ST2 a Novel Biomarker of Intracerebral Hemorrhage? Neurology. 2023;100:599–600. DOI: 10.1212/wnl.0000000000206861.

